# Cross-Cohort Evaluation of NanoString nCounter Data for Recurrence Prediction in Colorectal Cancer

**DOI:** 10.64898/2026.08.13.26360359

**Authors:** Puck Quarles van Ufford, Rasmus Dahlin Bojesen, Lars Rønn Olsen, Ismail Gögenur, Ole Lund

## Abstract

Gene expression-based prognostic models have shown promise for predicting recurrence in colorectal cancer (CRC), but their clinical implementation remains limited. The NanoString nCounter platform provides a practical alternative to RNA sequencing and microarrays through standardized, cost-effective gene expression profiling that is compatible with routine clinical samples. In this study, we evaluated whether NanoString nCounter gene expression data improve prediction of recurrence following curative CRC surgery.

Gene expression profiles from the NanoString PanCancer IO 360™ panel were analyzed in two independent CRC cohorts (cohort A, n = 189; cohort B, n = 131). Differential gene expression analyses and Cox proportional hazards models were used to assess the prognostic value of gene expression alone and in combination with established clinical risk factors. Model performance was evaluated by five-fold cross-validation and external validation between cohorts using the concordance index (C-index) and Kaplan-Meier risk stratification.

The two cohorts differed significantly in recurrence-free survival, and differential expression analysis demonstrated marked cohort-specific transcriptional patterns. Ninety-one recurrence-associated genes were identified in cohort A, whereas no significant genes were detected in cohort B, with poor agreement in gene-level differential expression between cohorts (Pearson r = 0.128). Across all prediction models, external performance was modest, and inclusion of gene expression data did not improve prediction beyond clinical variables. The clinical baseline model, incorporating age, UICC stage, and tumor site, consistently achieved the highest cross-cohort performance, with UICC stage emerging as the strongest predictor of recurrence. Although overall discrimination was moderate, the baseline model successfully stratified patients into significantly different high- and low-risk groups across cohorts.

These findings indicate that prognostic gene expression signatures derived from NanoString data showed limited reproducibility across independent cohorts and provided little additional predictive value beyond established clinical factors. The results highlight the importance of external validation and suggest that robust clinical variables remain the most reliable predictors of recurrence risk in this setting.

## Introduction

Colorectal cancer (CRC) is a highly heterogeneous disease, with substantial variation in molecular characteristics and clinical outcomes (1–3). Despite advances in prognostic modeling, risk stratification to predict patient outcomes remains challenging (4,5). Disease recurrence is one particularly clinically important outcome, which occurs in approximately 20-30% of stage I-III CRC patients, and even more frequently in patients with metastatic disease (6,7). Improved recurrence risk stratification is highly relevant, especially as the development of individually tailored post-operative surveillance strategies remains an active area of research (8,9).

Substantial efforts have been made in addressing CRC heterogeneity, including molecular classification based on gene expression, most notably the consensus molecular subtype (CMS) framework (10,11). Gene expression profiling has also been widely used in the development of prognostic prediction models (9,12). Most transcriptomics studies in CRC have relied on microarray or RNA sequencing (RNA-seq) technologies. While these methods enable comprehensive transcriptomic profiling, its clinical implementation can be impeded by cost, infrastructure requirements, longer turnaround time, the need for specialized expertise, and reliance on fresh frozen samples (4,9,13–16). Consequently, despite promising results, the integration of gene expression-based models into routine clinical practice remains limited.

The NanoString nCounter platform offers a potential alternative for clinical gene expression profiling. This technology provides several advantages, including lower costs, simplified workflows, faster processing times, and robust performance with formalin-fixed,paraffin-embedded (FFPE) tissue samples (17–19). These characteristics make the NanoString assay particularly attractive for translational applications. Several custom NanoString gene panels have already been developed for CRC profiling (16,20–25). However, commercially available panels such as the nCounter PanCancer IO 360™ panel offer practical advantages, including standardized design and easier implementation across hospitals.

In this study, we investigate whether gene expression data from the NanoString nCounter PanCancer IO 360™ panel can be used to predict CRC recurrence. We first establish a baseline prediction model based on established clinical risk factors. We then explore the prognostic value of the gene expression data, specifically by including differentially expressed genes, the full panel, and genes selected through univariate Cox regression. The models were trained on two independent patient cohorts. For the best-performing models, the cross-validation predictions were consolidated into a final risk classification of the patients in their respective test sets as high- or low-risk, using majority voting based on the median risk score predicted on the training data.

## Methods

### Data

NanoString nCounter gene expression data and clinical data were obtained from two independent cohorts at Slagelse Hospital, Denmark. Cohort A included 220 CRC patients, and cohort B (26) included 140 patients.

For each patient, one .RCC file containing raw NanoString nCounter data was available. The data were generated from formalin-fixed paraffin-embedded (FFPE) tumor tissue samples collected during CRC resection surgery. The samples were previously analyzed using the NanoString nCounter PanCancer IO 360™ Panel (NanoString Technologies, Seattle, WA, USA), which includes 770 gene probes, targeting 750 endogenous genes, 20 housekeeping genes, as well as 8 negative control probes, and 6 positive control probes.

Only patients with stage I-III CRC were included in the study. The survival outcomes used in the study were overall survival (OS), defined as the time from surgery to death, and recurrence-free survival (RFS), defined as the time from the surgery to either recurrence or death.

### Preparation of NanoString Data

NanoString nCounter data were preprocessed separately for each cohort. Raw expression data and associated metadata were extracted from .RCC files using the NanoStringQCPro R package (v1.30.0) (27). Quality control was performed in accordance with NanoString Technologies guidelines (28). Imaging quality was assessed by requiring that the proportion of counted fields of view (FOV) exceeded 75% of available FOV. Binding density was required to fall within the recommended range of 0.1-2.25. Linearity of the positive control probes was evaluated using the coefficient of determination (R^2^ > 0.95). The limit of detection was assessed by requiring that the POS_E positive control probe count exceeded the mean of the negative control probes by at least two standard deviations. Samples failing any of these criteria were excluded from further analysis. Following quality control, expression values were normalized using the nSolver-based normalization workflow implemented in the NanoTube R package (v1.4.0) (29).

### Batch Correction

Normalized expression values were log-transformed prior to batch correction. Gene-wise standardization was performed across cohorts to account for batch effects. For each gene, the mean and standard deviation were estimated separately within each cohort, and expression values were centered and scaled relative to the cohort-specific distribution. The standardized values were subsequently rescaled using the overall gene-level standard deviation across cohorts to maintain comparable variance across genes.

Principal component analysis (PCA) was then performed on the scaled expression data to identify and exclude outlier samples. Outliers were defined as samples with an absolute Z-score greater than 3 on either of the first two principal components.

### Splitting the Data

Each cohort was randomly split into a training set (80%) and a test set (20%), while maintaining the OS and RFS rate. The training set was used for differential gene expression analysis and model training. Model performance was evaluated both on the internal held-out test set and through cross-cohort external validation, where models trained in one cohort were tested on the full independent cohort.

### Differential Gene Expression Analysis

Differential gene expression analysis was performed using the DESeq2 R package (v1.38.3)(30) on the raw expression data. Within each cohort, patients who experienced cancer recurrence were compared with those who did not. Log2 fold changes were estimated using the non-recurrence group as the reference. Statistical significance was assessed using Wald tests, and p-values were adjusted for multiple testing using the Benjamini-Hochberg procedure. Differentially expressed genes (DEGs) were defined as genes with an adjusted p-value < 0.05.

### Survival Analysis

Survival models were trained using five-fold cross-validation. Cox proportional hazards models were fitted using the survival R package (v3.4.0) (31). For each cohort, four models were constructed: (1) the baseline model, which included established recurrence risk factors, namely age >75 years, UICC cancer stage, and primary tumor site (rectum vs. colon); (2) the DEG model, which extended the baseline model by including the DEGs identified in the differential expression analysis of the corresponding cohort; (3) the full panel model, which included the baseline features together with all 750 endogenous genes from the NanoString panel; and (4) the Cox-selected model, with included the baseline features and genes that were significant (p < 0.05) in univariate Cox regression analyses. The model performance was evaluated using the concordance index (C-index).

To further investigate the ability of the best-performing models to stratify patients according to recurrence risk, the five cross-validation models were used to assign patients in the test set to either a high-risk or low-risk group. The risk groups were based on the median risk score calculated in the training data. Final classification was determined by majority voting across the five models. Kaplan-Meier analysis was subsequently used to evaluate the separation between the risk groups with respect to RFS.

## Results

### Cohort Characteristics and Data Preprocessing

A total of 194 patients in cohort A and 137 patients in cohort B were initially identified with stage I-III CRC. Two patients from cohort A were excluded due to missing recurrence status, and one additional patient was excluded from cohort A because the sample failed to meet the limit-of-detection threshold. After quality control, 191 patients remained in cohort A and all 137 patients remained in cohort B.

Gene expression data were preprocessed separately for each cohort. Normalization improved the overall structure of the data, as evidenced by log-transformed expression distributions that more closely approximate a normal distribution, and relative log expression values that were more evenly distributed across samples and centered around zero (Fig. S1-S2).

Batch correction was subsequently applied to reduce systematic differences between the cohorts. Prior to correction, expression values differed significantly between cohorts (Student’s t-test, p < 2.2 × 10^-16^), and samples clustered by cohort in principal component analysis (PCA) space. Despite this clear separation, the mean gene expression across cohorts was highly correlated (Pearson r = 0.968) (Fig. S3A-C). After batch correction, differences in the expression distribution were substantially reduced but remained statistically significant (Student’s t-test, p = 0.0002). PCA demonstrated considerable overlap between the cohorts, although several strong outliers remained. The correlation in mean gene expression between cohorts decreased (Pearson r = 0.801) (Fig. S3D-F).

Two PCA outliers from cohort A and six from cohort B were subsequently excluded. The expression distributions were no longer significantly different between cohorts (Student’s t-test, p = 0.961), and samples from both cohorts largely overlapped in PCA space. The correlation between the mean gene expression partially recovered (Pearson r = 0.863) (Fig. S3G-I). The final dataset comprised 189 patients in cohort A and 131 patients in cohort B.

Baseline clinical and demographic characteristics were summarized in Table 1. Within each cohort, patients were stratified by recurrence status. UICC cancer stage differed significantly between recurrence groups in both cohorts, whereas sex, age, and primary tumor site were comparable between groups. Comparisons between cohorts revealed significant differences in age (p = 0.008), cancer stage (p = 0.005), and primary tumor site (p < 0.001).

**Table 1.** Cohort characteristics stratified by recurrence status in cohorts A and B. Baseline characteristics are shown separately for cohorts A and B and stratified by recurrence status (recurrence vs. no recurrence). P values indicate comparisons between patients with and without recurrence within each cohort.

| Feature | Cohort A |  |  | Cohort B |  |  |
| --- | --- | --- | --- | --- | --- | --- |
|  | Recurrence | No recurrence | p-value | Recurrence | No recurrence | p-value |
| n | 32 | 157 |  | 19 | 112 |  |
| Sex = male (%) | 20 (62.5) | 79 (50.3) | 0.288 | 12 (63.2) | 62 (55.4) | 0.701 |
| Age (mean (SD)) | 70.0 (10.0) | 71.2 (10.3) | 0.526 | 66.7 (7.5) | 68.3 (9.0) | 0.474 |
| Age > 75 (%) | 13 (40.6) | 57 (36.3) | 0.795 | 1 (5.3) | 22 (19.6) | 0.231 |
| UICC cancer stage (%) |  |  | < 0.001 |  |  | 0.006 |
| 1 | 2 (6.2) | 29 (18.5) |  | 2 (10.5) | 31 (27.7) |  |
| 2 | 9 (28.1) | 90 (57.3) |  | 3 (15.8) | 42 (37.5) |  |
| 3 | 21 (65.6) | 38 (24.2) |  | 14 (73.7) | 39 (34.8) |  |
| Primary site = rectum (%) | 2 (6.2) | 6 (3.9) | 0.900 | 6 (31.6) | 39 (34.8) | 0.989 |

### Survival Outcomes Differ Between Cohorts

Overall survival (OS) (Fig. 1A) and recurrence-free survival (RFS) (Fig. 1B) differed significantly between cohorts A and B. Kaplan-Meier analysis showed poorer survival outcomes in cohort A for both endpoints, and log-rank tests confirmed statistically significant differences in OS (p = 0.007) and RFS (p = 0.043). The median follow-up time was 3.78 years in cohort A and 4.69 years in cohort B. During follow-up, 43 patients (22.8%) in cohort A died, compared with 17 patients (13.5%) in cohort B. Disease recurrence occurred in 32 patients (16.9%) in cohort A and in 19 patients (14.5%) in cohort B.

**Figure 1.**
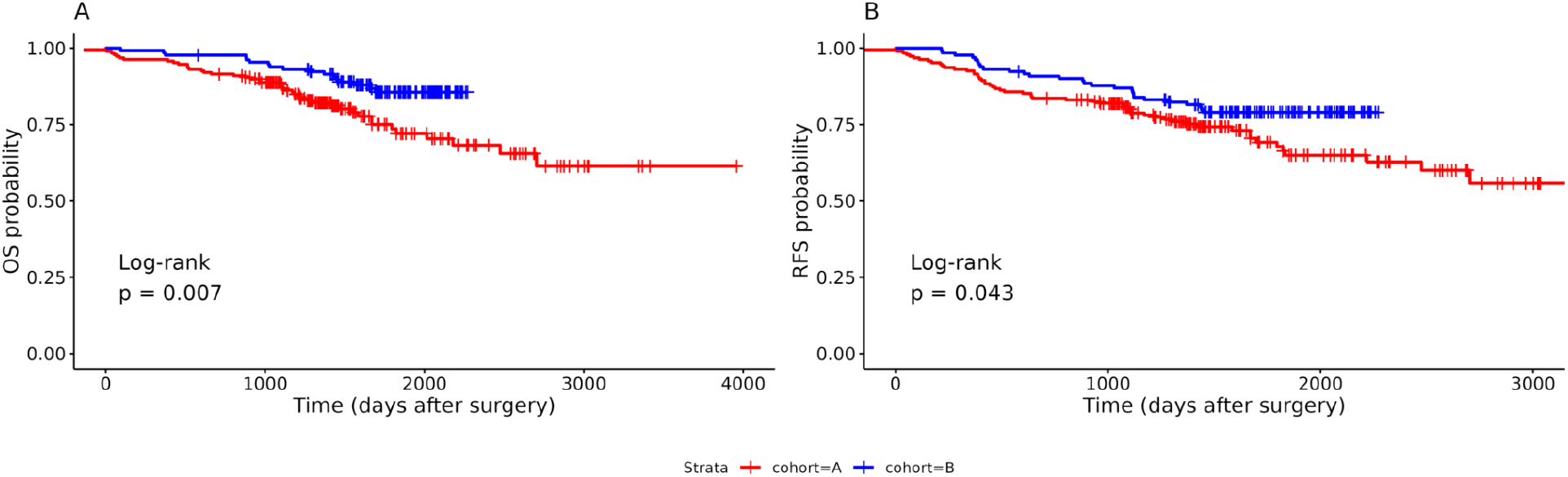
Kaplan-Meier survival curves for cohorts A and B. Kaplan-Meier plots for (A) overall survival (OS) and (B) recurrence-free survival (RFS) for cohort A (red) and cohort B (blue). P values from log-rank tests indicate significant differences between the two cohorts for both outcomes.

### Differential Gene Expression is Cohort-Specific

Differential gene expression analysis revealed distinct patterns between cohort A and cohort B (Fig. 2A-B). In cohort A, 91 genes were significantly differentially expressed (Benjamini-Hochberg adjusted p < 0.05), when comparing patients with and without recurrence. In contrast, no genes met the same significance threshold in cohort B. Importantly, comparison of log2 fold changes across all genes revealed low concordance between cohorts (Pearson r = 0.128), indicating substantial cohort-specific transcriptional patterns (Fig. 2C).

**Figure 2.**
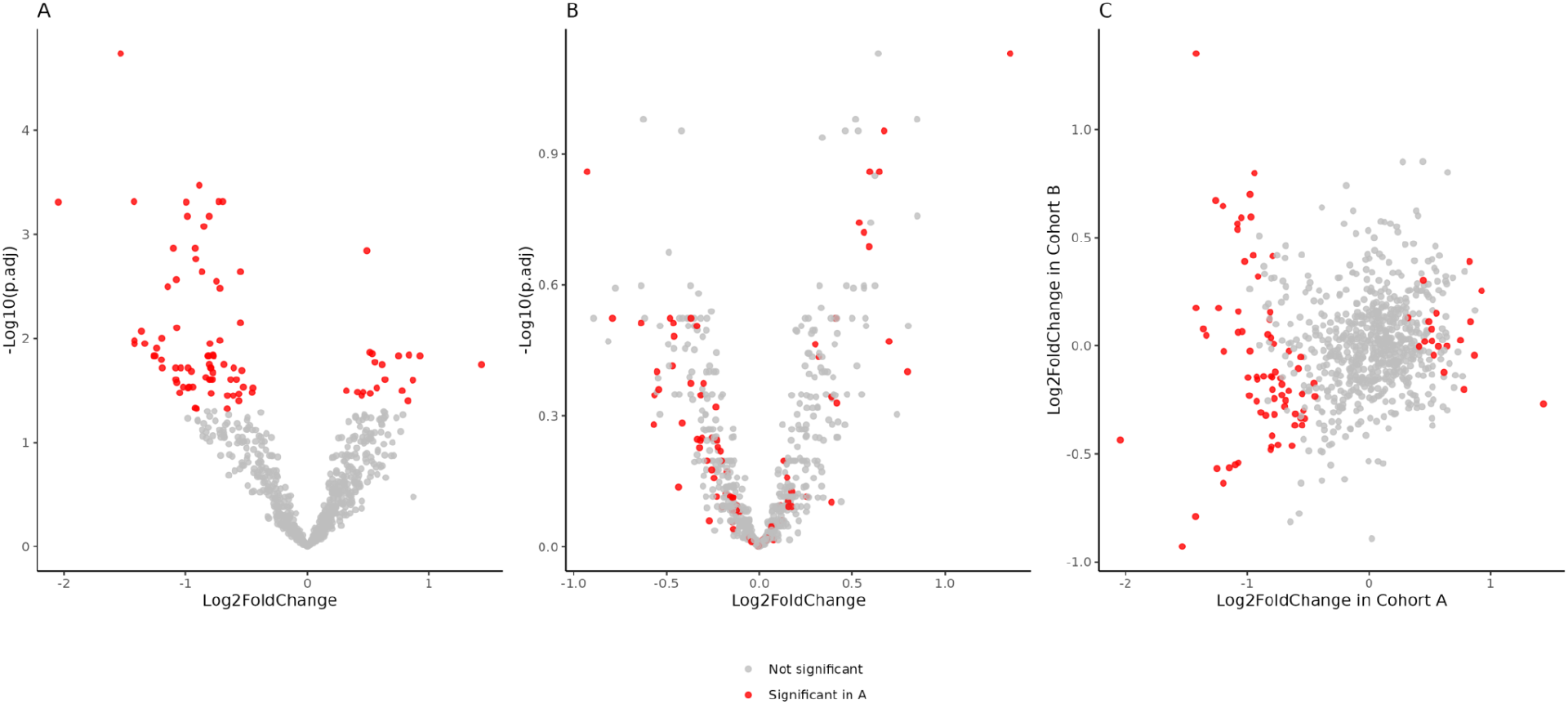
Differential gene expression analysis in cohorts A and B. Volcano plots showing differential gene expression results for (A) cohort A and (B) cohort B. (C) Scatter plot of log fold change values for each gene comparing cohort A and cohort B. Genes differentially expressed in cohort A are shown in red and non-significant genes in grey. No differentially expressed genes were identified in cohort B.

### Cox Proportional Hazards Models Demonstrate Moderate Predictive Performance for Recurrence-Free Survival

The four Cox proportional hazards models described in the Methods were trained separately in each cohort. Model performance was first evaluated using the held-out internal test sets and subsequently assessed in the other cohort to evaluate external generalizability.

Across all models, predictive performance did not generalize well between cohorts. In both cases, the highest concordance index (C-index) in the external test set was achieved by the baseline model (Table 2).

**Table 2.** Performance of Cox proportional hazards models trained in cohorts A and B. Concordance indices (C-index) with standard deviations for four Cox proportional hazards models trained separately in cohorts A and B. The baseline model included age, tumor site, and cancer stage. Model 2 additionally included differentially expressed genes (DEGs), Model 3 included all genes from the nCounter PanCancer IO 360 Panel, and Model 4 included genes selected by univariate Cox analysis in addition to the baseline variables. For each training cohort, columns report the C-index in the training set, the internal test set, and external validation in the other cohort.

| Model | Trained on cohort A |  |  | Trained on cohort B |  |  |
| --- | --- | --- | --- | --- | --- | --- |
|  | C-index in training set | C-index in test set | C-index in cohort B | C-index in training set | C-index in test set | C-index in cohort A |
| Baseline: advanced age + tumor site + cancer stage | 0.622 (0.07) | 0.650 (0.02) | 0.590 (0.01) | 0.573 (0.09) | 0.451 (0.03) | 0.570 (0.03) |
| Baseline + DEGs | 0.509 (0.12) | 0.538 (0.08) | 0.468 (0.04) | - | - | - |
| Baseline + all panel genes | 0.547 (0.10) | 0.511 (0.04) | 0.486 (0.02) | 0.470 (0.14) | 0.541 (0.24) | 0.496 (0.03) |
| Baseline + Cox-selected genes | 0.558 (0.13) | 0.499 (0.04) | 0.537 (0.01) | 0.457 (0.20) | 0.532 (0.17) | 0.499 (0.054) |

To further investigate the drivers of prediction in the baseline model, the coefficients from the cross-validation models were examined (Fig. 3). In both cohorts, UICC stage emerged as the strongest predictor of recurrence-free survival. In cohort A, however, primary tumor site showed the largest average effect size across models.

**Figure 3.**
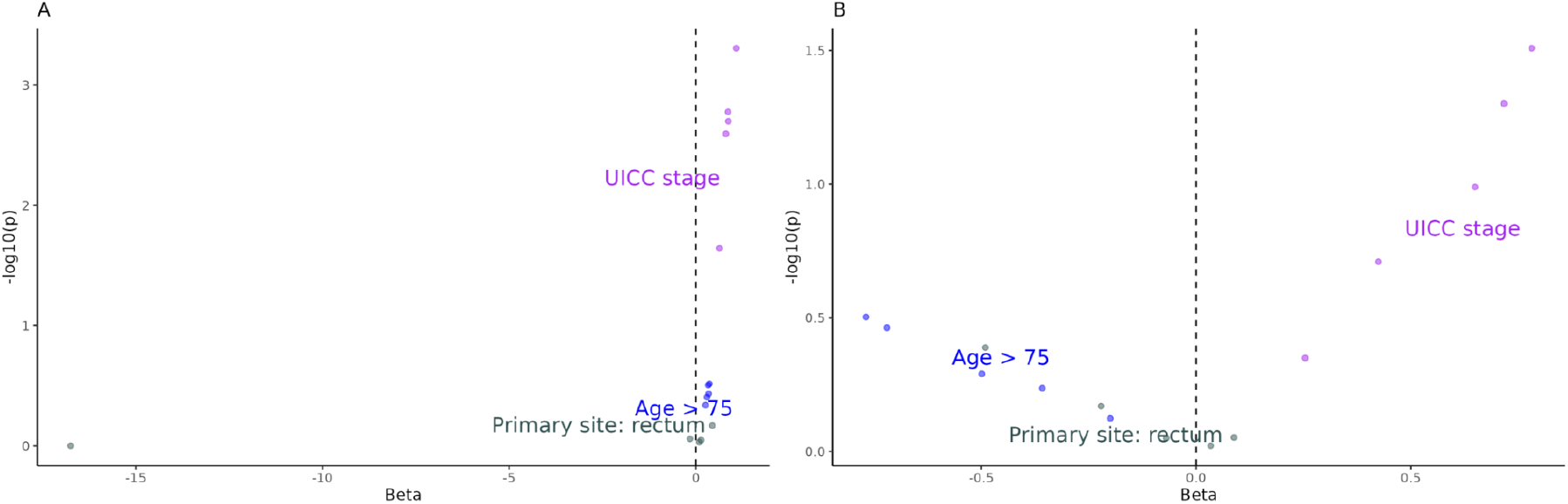
Coefficients of features included in baseline models. Scatter plot of the coefficients and p-values of the baseline model trained on cohort A (A) and cohort B (B) across five cross-validation models.

### Majority-Voting Separates High-Risk and Low-Risk Patients

To further evaluate the ability of the baseline model to stratify patients by recurrence risk, the five cross-validation models were used to classify patients in each test set into high- or low-risk groups based on the median risk score calculated in the training data. Final classification was determined by majority voting across the five models.

Kaplan-Meier analysis showed that neither model significantly discriminated between high- and low-risk groups in the internal test sets, likely due to the limited number of patients (cohort A: n = 38; cohort B: n = 26). However, both models achieved significant risk stratification when applied to the independent cohort. The baseline model trained in cohort A significantly separated risk groups in cohort B (p = 0.04), with a 2.05-fold higher 5-year risk in the high-risk group compared with the low-risk group. Similarly, the baseline model trained in cohort B significantly stratified patients in cohort A (p < 0.0001), with a 2.24-fold higher 5-year risk in the high-risk group.

**Figure 4.**
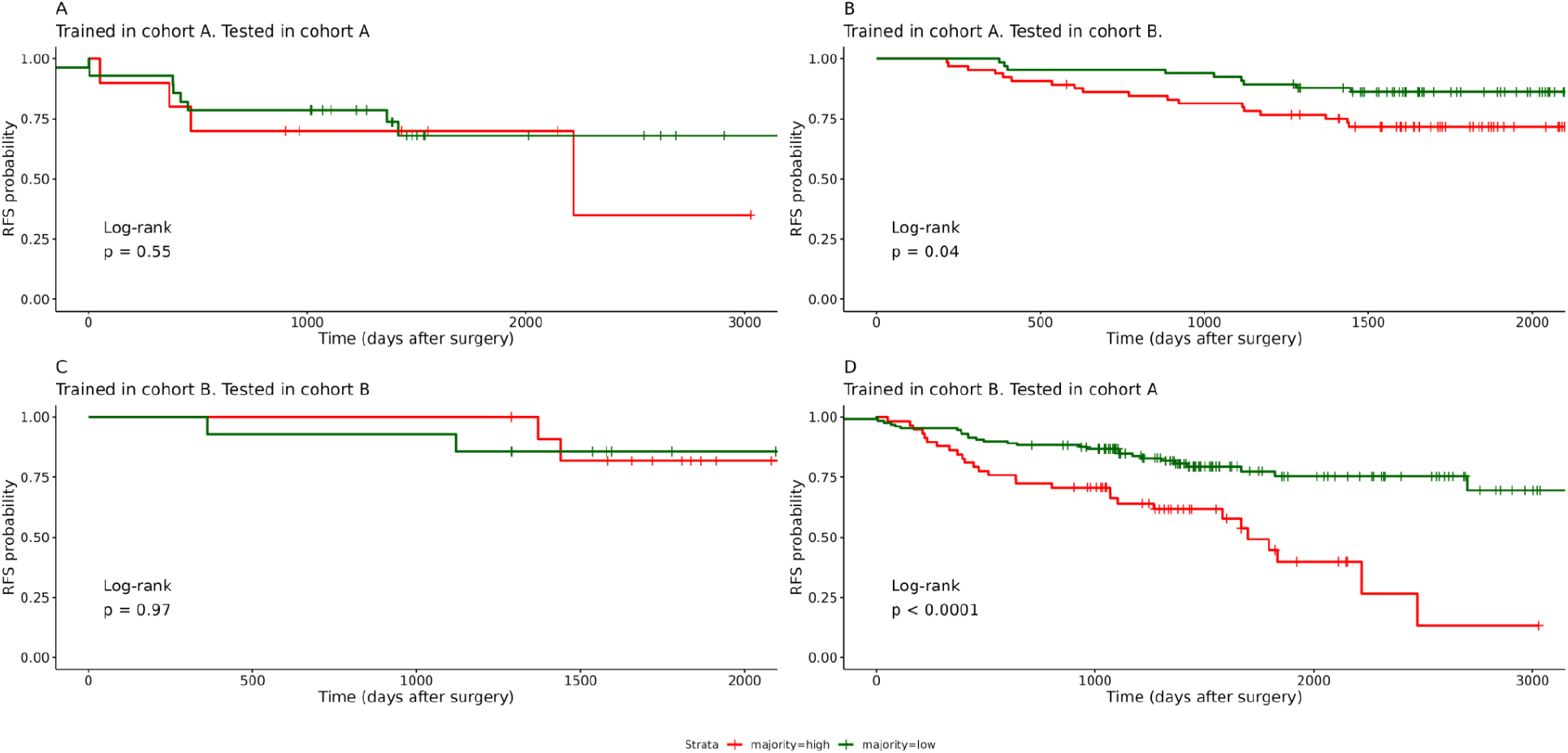
Kaplan-Meier analysis based on majority-vote risk classification across five cross-validation models. Kaplan-Meier curves comparing high- and low-risk groups defined by majority voting across five-fold cross-validation Cox models. (A-B) Baseline model trained in cohort A and tested in the internal test set (A) and in cohort B (B). (C-D) Baseline model trained in cohort B and tested in the internal test set (C) and in cohort A (D).

## Discussion

In this study, we aimed to predict recurrence-free survival after CRC surgery using NanoString nCounter expression data. First, we compared the gene expression profiles between patients with and without recurrence in order to identify significantly differentially expressed genes. Here, we identified a strong difference between the results of the two cohorts. The variability between the cohorts most likely arises from clinical differences. The limited agreement in differential gene expression patterns between the two cohorts may further highlight the challenges associated with identifying robust prognostic gene signatures. Our findings underscore the importance of evaluating candidate gene expression signatures in independent cohorts and using external validation when developing prognostic models.

We next trained Cox proportional hazards models using baseline predictors including age, cancer stage, and tumor site. We then evaluated whether incorporating gene expression data improved predictive performance by testing DEGs, the full nCounter panel, and genes selected through univariate Cox regression. Overall, the survival models demonstrated limited ability to rank patients according to recurrence-free survival, as indicated by the C-index. In both cohorts, the baseline model achieved the best performance in the external test sets.

Despite the modest C-index values, stratification of patients into high- and low-risk groups revealed significant differences in outcomes, with clinically meaningful differences in 5-year recurrence risk between the two groups. This suggests that, although the models showed limited discrimination when evaluated by the C-index, they may still provide practical value for patient stratification. One explanation for the modest C-index values is that the metric evaluates the ability to rank individual patients across the entire survival distribution, which can be difficult in relatively small cohorts with moderate event rates. In contrast, risk-group stratification focuses on identifying subsets of patients with clearly different prognoses, which may be more relevant for clinical decision-making, such as identifying patients who may benefit from intensified post-operative surveillance.

There are several limitations to this study. First of all, we were able to identify a significant batch effect between the cohorts, both in terms of expression data and in terms of clinical outcomes. The difference in expression data is addressed through data normalization and batch correction; however, there is no clear consensus in the literature on how to preprocess NanoString nCounter data. Here, we adhered to the in-house guidelines by NanoString to do quality control and we used NanoString nSolver methods for normalization, using the control probes and housekeeping genes. Still, several other methods have been developed for NanoString nCounter normalization (19,32–37), which may have led to different results.

Another limitation of the study is that both cohorts were generated in the same hospital and the patients included were from a relatively homogenous Danish background. We aimed to evaluate the generalizability of the results by working with two distinct, independent cohorts; however, there may still be limitations to the generalizability in other contexts.

## Data Availability

All data produced in the present study are available upon reasonable request to the authors.

## Referrences

1. Molinari C, Marisi G, Passardi A, Matteucci L, De Maio G, Ulivi P. Heterogeneity in Colorectal Cancer: A Challenge for Personalized Medicine? Int J Mol Sci. 2018 Dec;19(12):12. doi:10.3390/ijms19123733

2. Saoudi González N, Salvà F, Ros J, Baraibar I, Rodríguez-Castells M, García A, et al. Unravelling the Complexity of Colorectal Cancer: Heterogeneity, Clonal Evolution, and Clinical Implications. Cancers. 2023 Jan;15(16):16. doi:10.3390/cancers15164020

3. Sobral D, Martins M, Kaplan S, Golkaram M, Salmans M, Khan N, et al. Genetic and microenvironmental intra-tumor heterogeneity impacts colorectal cancer evolution and metastatic development. Commun Biol. 2022 Sep 9;5(1):1–14. doi:10.1038/s42003-022-03884-x

4. Koncina E, Haan S, Rauh S, Letellier E. Prognostic and Predictive Molecular Biomarkers for Colorectal Cancer: Updates and Challenges. Cancers. 2020 Jan 30;12(2). doi:10.3390/cancers12020319

5. Goh V, Mallett S, Rodriguez-Justo M, Boulter V, Glynne-Jones R, Khan S, et al. Evaluation of prognostic models to improve prediction of metastasis in patients following potentially curative treatment for primary colorectal cancer: the PROSPECT trial. Health Technol Assess. 2025 Apr 10;29(8):1–91. doi:10.3310/BTMT7049

6. van der Stok EP, Spaander MCW, Grünhagen DJ, Verhoef C, Kuipers EJ. Surveillance after curative treatment for colorectal cancer. Nat Rev Clin Oncol. 2017 May;14(5):297–315. doi:10.1038/nrclinonc.2016.199 PubMed PMID: 27995949.

7. Qaderi SM, Galjart B, Verhoef C, Slooter GD, Koopman M, Verhoeven RHA, et al. Disease recurrence after colorectal cancer surgery in the modern era: a population-based study. Int J Colorectal Dis. 2021 Nov 1;36(11):2399–410. doi:10.1007/s00384-021-03914-w

8. Nors J, Iversen LH, Erichsen R, Gotschalck KA, Andersen CL. Incidence of Recurrence and Time to Recurrence in Stage I to III Colorectal Cancer. JAMA Oncol. 2024 Jan;10(1):54–62. doi:10.1001/jamaoncol.2023.5098 PubMed PMID: 37971197; PubMed Central PMCID: PMC10654928.

9. Ahluwalia P, Kolhe R, Gahlay GK. The clinical relevance of gene expression based prognostic signatures in colorectal cancer. Biochim Biophys Acta BBA - Rev Cancer. 2021 Apr 1;1875(2):188513. doi:10.1016/j.bbcan.2021.188513

10. Guinney J, Dienstmann R, Wang X, de Reyniès A, Schlicker A, Soneson C, et al. The consensus molecular subtypes of colorectal cancer. Nat Med. 2015 Nov;21(11):1350–6. doi:10.1038/nm.3967

11. Dunne PD, Arends MJ. Molecular pathological classification of colorectal cancer—an update. Virchows Arch. 2024;484(2):273–85. doi:10.1007/s00428-024-03746-3 PubMed PMID: 38319359; PubMed Central PMCID: PMC10948573.

12. Saadh MJ, Allela OQB, Kareem RA, Baldaniya L, Ballal S, Vashishth R, et al. Prognostic gene expression profile of colorectal cancer. Gene. 2025 Jul 5;955:149433. doi:10.1016/j.gene.2025.149433 PubMed PMID: 40122415.

13. Bayle A, Bonastre J, Chaltiel D, Latino N, Rouleau E, Peters S, et al. ESMO study on the availability and accessibility of biomolecular technologies in oncology in Europe. Ann Oncol. 2023 Oct 1;34(10):934–45. doi:10.1016/j.annonc.2023.06.011

14. Ding X, Huang H, Fang Z, Jiang J. From Subtypes to Solutions: Integrating CMS Classification with Precision Therapeutics in Colorectal Cancer. Curr Treat Options Oncol. 2024 Dec;25(12):1580–93. doi:10.1007/s11864-024-01282-5 PubMed PMID: 39589648.

15. Włodarczyk M, Włodarczyk J, Siwiński P, Sobolewska-Włodarczyk A, Fichna J. Genetic Molecular Subtypes in Optimizing Personalized Therapy for Metastatic Colorectal Cancer. Curr Drug Targets. 2018;19(15):1731–7. doi:10.2174/1389450119666180803122744

16. Chen X, Deane NG, Lewis KB, Li J, Zhu J, Washington MK, et al. Comparison of Nanostring nCounter® Data on FFPE Colon Cancer Samples and Affymetrix Microarray Data on Matched Frozen Tissues. PLOS ONE. 2016 May 13;11(5):e0153784. doi:10.1371/journal.pone.0153784

17. Eastel JM, Lam, Ka Wai, Lee, Nga Lam, Lok, Wing Yan, Tsang, Andy Hin Fung, Pei, Xiao Meng, et al. Application of NanoString technologies in companion diagnostic development. Expert Rev Mol Diagn. 2019 Jul 3;19(7):591–8. doi:10.1080/14737159.2019.1623672 PubMed PMID: 31164012.

18. Geiss GK, Bumgarner RE, Birditt B, Dahl T, Dowidar N, Dunaway DL, et al. Direct multiplexed measurement of gene expression with color-coded probe pairs. Nat Biotechnol. 2008 Mar;26(3):317–25. doi:10.1038/nbt1385

19. Waggott D, Chu K, Yin S, Wouters BG, Liu FF, Boutros PC. NanoStringNorm: an extensible R package for the pre-processing of NanoString mRNA and miRNA data. Bioinformatics. 2012 Jun 1;28(11):1546–8. doi:10.1093/bioinformatics/bts188

20. Ragulan C, Eason K, Fontana E, Nyamundanda G, Tarazona N, Patil Y, et al. Analytical Validation of Multiplex Biomarker Assay to Stratify Colorectal Cancer into Molecular Subtypes. Sci Rep. 2019 May 21;9(1):7665. doi:10.1038/s41598-019-43492-0

21. Marisa L, Blum Y, Taieb J, Ayadi M, Pilati C, Le Malicot K, et al. Intratumor CMS Heterogeneity Impacts Patient Prognosis in Localized Colon Cancer. Clin Cancer Res. 2021 Sep 1;27(17):4768–80. doi:10.1158/1078-0432.CCR-21-0529

22. Morris JS, Luthra R, Liu Y, Duose DY, Lee W, Reddy NG, et al. Development and Validation of a Gene Signature Classifier for Consensus Molecular Subtyping of Colorectal Carcinoma in a CLIA-Certified Setting. Clin Cancer Res. 2021 Jan 4;27(1):120–30. doi:10.1158/1078-0432.CCR-20-2403

23. Piskol R, Huw L, Sergin I, Kljin C, Modrusan Z, Kim D, et al. A Clinically Applicable Gene-Expression Classifier Reveals Intrinsic and Extrinsic Contributions to Consensus Molecular Subtypes in Primary and Metastatic Colon Cancer. Clin Cancer Res. 2019 Jul 15;25(14):4431–42. doi:10.1158/1078-0432.CCR-18-3032

24. Lenz HJ, Ou FS, Venook AP, Hochster HS, Niedzwiecki D, Goldberg RM, et al. Impact of Consensus Molecular Subtype on Survival in Patients With Metastatic Colorectal Cancer: Results From CALGB/SWOG 80405 (Alliance). J Clin Oncol. 2019 Aug;37(22):1876–85. doi:10.1200/JCO.18.02258

25. Torang A, van de Weerd S, Lammers V, van Hooff S, van den Berg I, van den Bergh S, et al. NanoCMSer: a consensus molecular subtype stratification tool for fresh-frozen and paraffin-embedded colorectal cancer samples. Mol Oncol. 2025;19(5):1332–46. doi:10.1002/1878-0261.13781

26. Gögenur M, Balsevicius L, Jensen SØ, Øgaard N, Lyskjær I, Justesen TF, et al. Preoperative positive ctDNA analysis is associated with the tumor microenvironment, and the risk of recurrence in non-metastatic colorectal cancer. Npj Precis Oncol. 2026 Jan 21;10(1):76. doi:10.1038/s41698-026-01288-2

27. Nickles D, Sandmann T, Ziman R, Bourgon R. NanoStringQCPro: Quality metrics and data processing methods for NanoString mRNA gene expression data [Internet]. 2022. Available from: http://bioconductor.org/packages/NanoStringQCPro/ doi:10.18129/B9.bioc.NanoStringQCPro

28. Gene Expression Data Analysis Guidelines (MAN-C0011-04). NanoString Technologies Inc.; 2017.

29. Class CA, Lukan CJ, Bristow CA, Do KA. Easy NanoString nCounter data analysis with the NanoTube. Bioinformatics. 2023 Jan 1;39(1):btac762. doi:10.1093/bioinformatics/btac762

30. Love MI, Huber W, Anders S. Moderated estimation of fold change and dispersion for RNA-seq data with DESeq2. Genome Biol. 2014 Dec 5;15(12):550. doi:10.1186/s13059-014-0550-8

31. Therneau TM. A Package for Survival Analysis in R [Internet]. 2022. Available from: https://CRAN.R-projects.org/package=survival

32. Wang H, Horbinski C, Wu H, Liu Y, Sheng S, Liu J, et al. NanoStringDiff: a novel statistical method for differential expression analysis based on NanoString nCounter data. Nucleic Acids Res. 2016 Nov 16;44(20):e151. doi:10.1093/nar/gkw677 PubMed PMID: 27471031; PubMed Central PMCID: PMC5175344.

33. Canouil M, Bouland GA, Bonnefond A, Froguel P, ‘t Hart LM, Slieker RC. NACHO: an R package for quality control of NanoString nCounter data [Internet]. [cited 2026 Apr 28]. Available from: 10.1093/bioinformatics/btz647

34. Molania R, Gagnon-Bartsch JA, Dobrovic A, Speed TP. A new normalization for Nanostring nCounter gene expression data. Nucleic Acids Res. 2019 Jul 9;47(12):6073–83. doi:10.1093/nar/gkz433 PubMed PMID: 31114909; PubMed Central PMCID: PMC6614807.

35. Brumbaugh CD, Kim HJ, Giovacchini M, Pourmand N. NanoStriDE: normalization and differential expression analysis of NanoString nCounter data. BMC Bioinformatics. 2011 Dec 16;12:479. doi:10.1186/1471-2105-12-479 PubMed PMID: 22177214; PubMed Central PMCID: PMC3273488.

36. Bhattacharya A, Hamilton AM, Furberg H, Pietzak E, Purdue MP, Troester MA, et al. An approach for normalization and quality control for NanoString RNA expression data. Brief Bioinform. 2021 May 20;22(3):bbaa163. doi:10.1093/bib/bbaa163 PubMed PMID: 32789507; PubMed Central PMCID: PMC8138885.

37. Jia G, Wang X, Li Q, Lu W, Tang X, Wistuba I, et al. RCRnorm: An integrated system of random-coefficient hierarchical regression models for normalizing NanoString nCounter data. Ann Appl Stat. 2019 Sep;13(3):1617–47. doi:10.1214/19-aoas1249 PubMed PMID: 33564347; PubMed Central PMCID: PMC7869841.

