## Supplementary material for "Cross-Cohort Evaluation of NanoString nCounter Data for Recurrence Prediction in Colorectal Cancer": Figure S

### Supplemental Materials

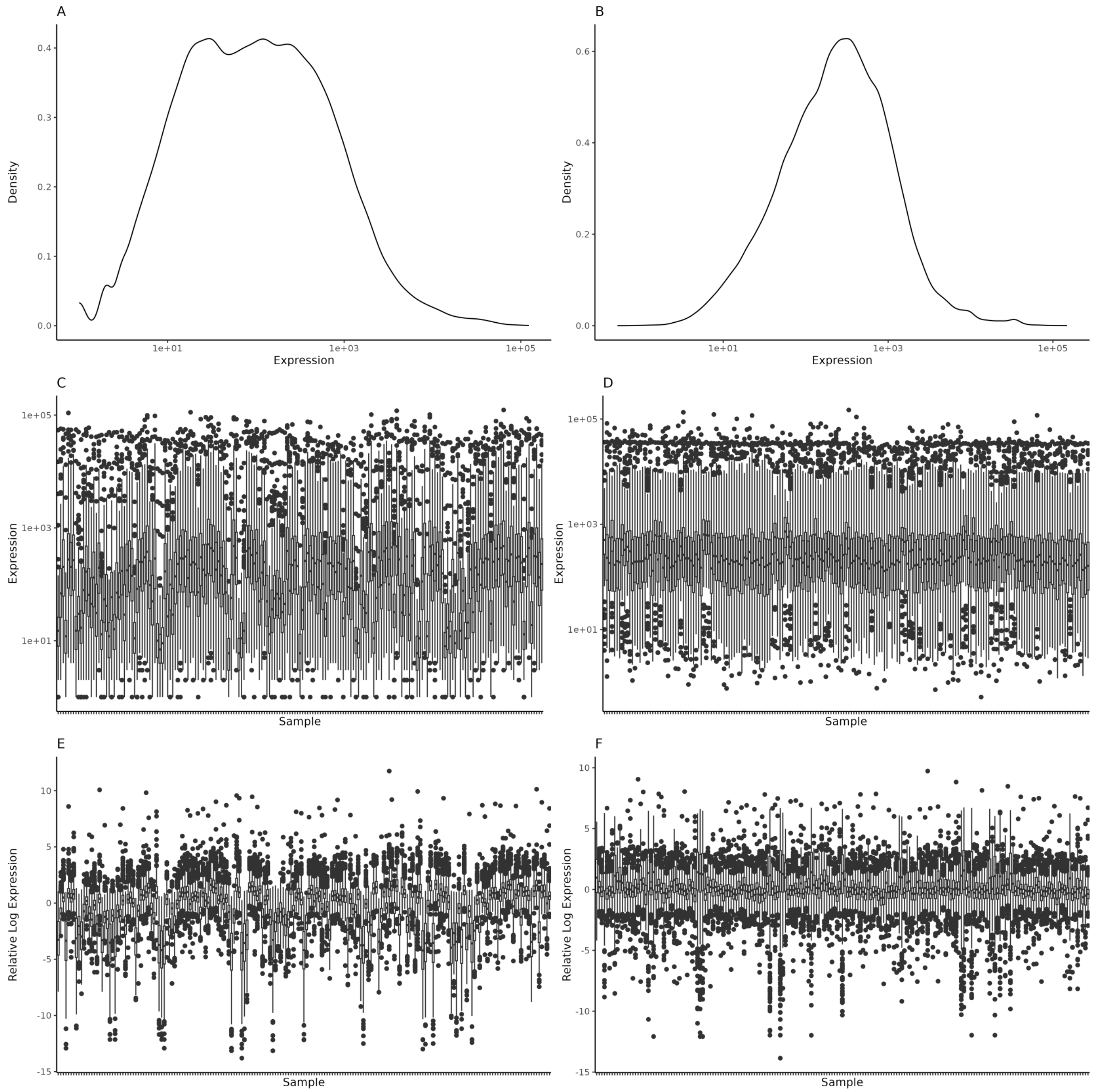


**Figure S1.** Normalization of gene expression data in cohort A.
Assessment of gene expression data before and after normalization in cohort A. (A, B) Density plots of log-scaled expression values pre- and post-normalization. (C, D) Box plots of log-scaled gene expression values per sample. (E, F) Relative log expression plots per sample.

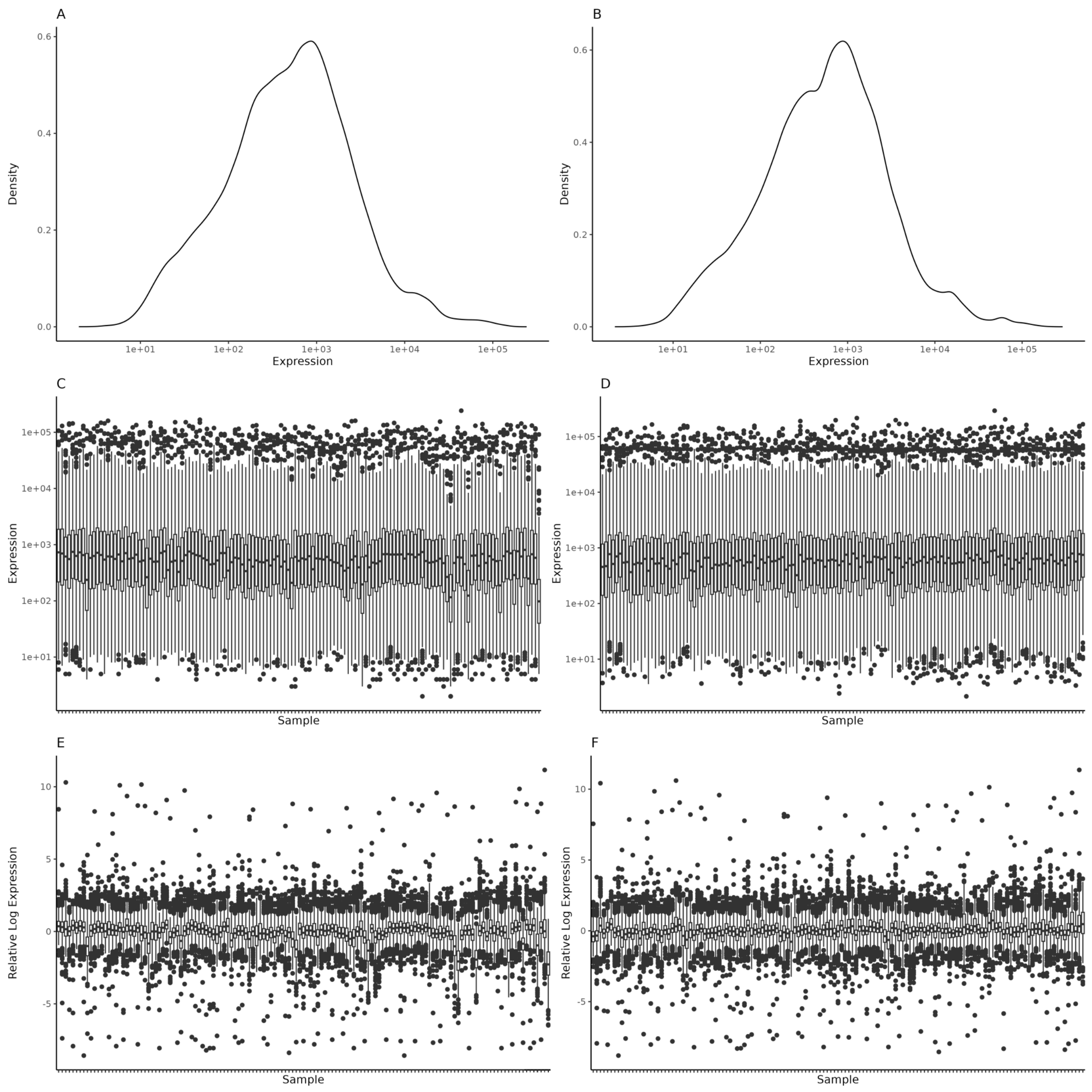


**Figure S2.** Normalization of gene expression data in cohort B.

Assessment of gene expression data before and after normalization in cohort B. (A, B) Density plots of log-scaled expression values pre- and post-normalization. (C, D) Box plots of log-scaled gene expression values per sample. (E, F) Relative log expression plots per sample.

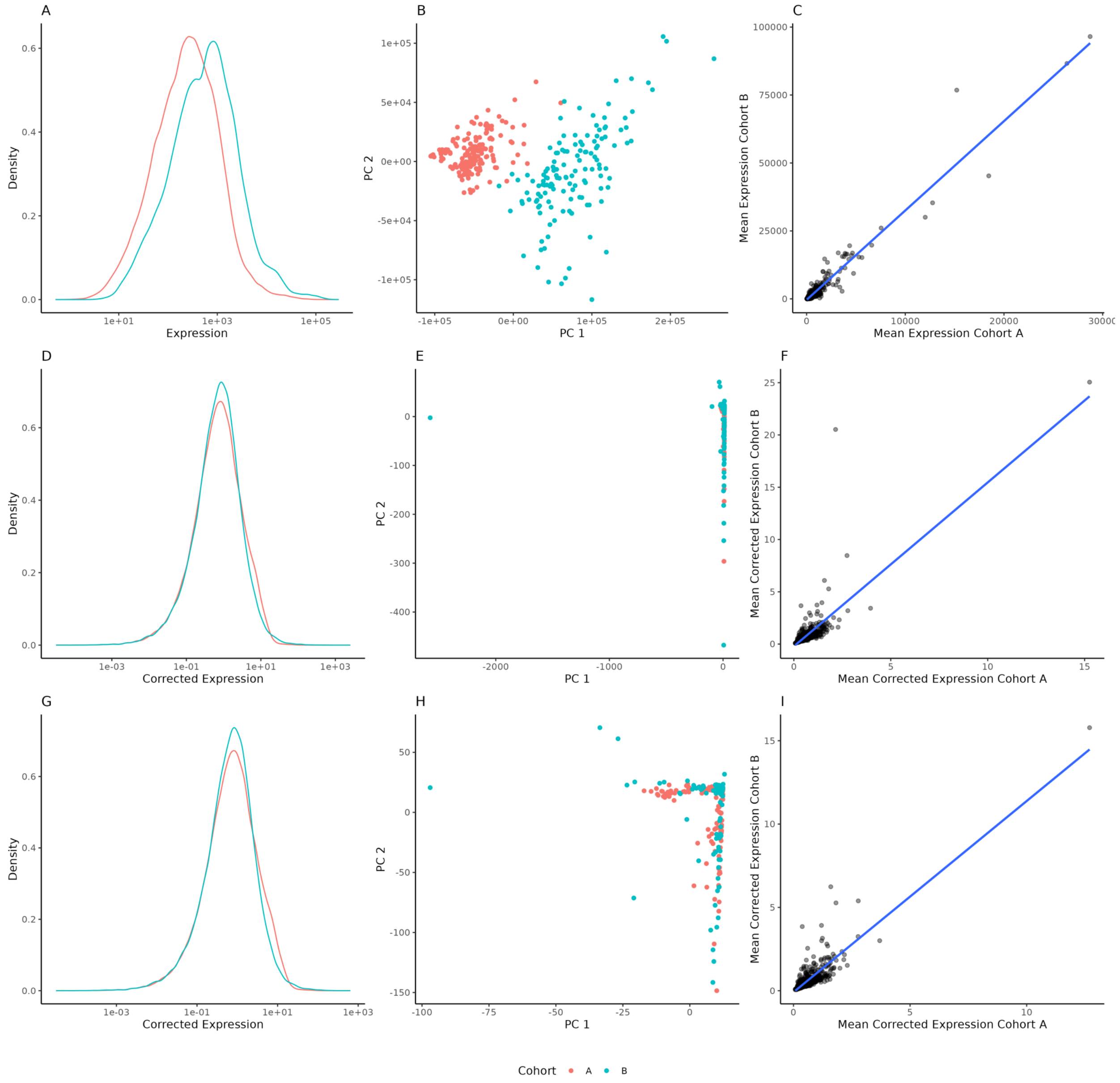


**Figure S3.** Batch correction and outlier removal of gene expression data.
Evaluation of preprocessing steps including batch correction and outlier removal. (A, D, G) Density plots of log-transformed gene expression values before correction, after batch correction, and after outlier removal, respectively. (B, E, H) Principal component analysis (PCA) plots. (C, F, I) Scatter plots comparing mean gene expression between cohorts A and B.
